# Geospatial assessment of the convergence of communicable and non-communicable diseases in South Africa

**DOI:** 10.1101/2023.03.01.23286636

**Authors:** Diego F. Cuadros, Claudia M. Moreno, Andrew Tomita, Urisha Singh, Stephen Olivier, Alison Castle, Yumna Moosa, Johnathan A Edwards, Hae-Young Kim, Mark J Siedner, Emily B Wong, Frank Tanser

## Abstract

**Background:** Several low- and middle-income countries are undergoing a rapid epidemiological transition with a rising burden of non-communicable diseases (NCDs). South Africa (SA) is a country with one of the largest HIV epidemics worldwide and a growing burden of NCDs where the collision of these epidemics poses a major public health challenge.

**Methods:** Using data from a large nationally representative survey, the South Africa Demographic and Health Survey (SADHS 2016), we conducted a geospatial analysis of several diseases including HIV, tuberculosis (TB), cardiovascular, respiratory, and metabolic diseases to identify areas with a high burden of co-morbidity within the country. We explored the spatial structure of each disease and the associations between diseases using different spatial and visual data methodologies. We also assessed the individual-level co-occurrence of HIV and the other diseases included in the analysis.

**Results:** The spatial distribution for HIV prevalence showed that this epidemic is most intense in the eastern region of the country, mostly within the Gauteng, Mpumalanga, and Kwazulu-Natal provinces. In contrast, chronic diseases had their highest prevalence rates in the southern region of the country, particularly in the Eastern and Western Cape provinces. Individual-level analyses were consistent with the spatial correlations and found no statistically significant associations between HIV infection and the presence of any NCDs.

**Conclusions:** We found no evidence of geospatial overlap between the HIV epidemic and NCDs in SA. These results evidence the complex epidemiological landscape of the country, characterized by geographically distinct areas exhibiting different health burdens. The detailed description of the heterogenous prevalence of HIV and NCDs in SA reported in this study could be a useful tool to inform and direct policies to enhance targeted health service delivery according to the local health needs of each community.

## BACKGROUND

Multimorbidity, defined by the World Health Organization (WHO) as the co-occurrence of two or more health conditions in the same individual and communities, is a growing global public health challenge (1, 2). The increase in multimorbidity worldwide is creating a high burden on individuals, healthcare systems, and societies (3–5). Multimorbidity increases mortality levels (6, 7), lowers the quality of life (8), and intensifies utilization of health services and associated costs (9). For many years, multimorbidity of non-communicable diseases (NCDs) was recognized as a burden affecting primarily high-income countries. In contrast, communicable diseases such as HIV and malaria have classically been the main burden for low/middle-income countries, and remain highly prevalent in much of the global south (10). However, modern sociodemographic and lifestyle changes have triggered a concomitant growing epidemic of NCDs in low/middle-income countries. The risk of multimorbidity of communicable and non-communicable diseases is, therefore, a unique challenge faced by low/middle-income countries, where healthcare resources are often limited (11–13).

Pre-existing epidemics of communicable diseases could impact the incidence, prevalence, and management of rising NCDs epidemics. Understanding the multimorbidity patterns, associations, and determinants could enhance the strategies developed to tackle the current health burdens in these countries.

Being the epicenter of the HIV and tuberculosis (TB) epidemics, sub-Saharan Africa (SSA) (10, 14) represents a region of interest to understand the collision of communicable disease and NCD epidemics. The effectiveness of modern antiretroviral therapy (ART) has changed the landscape of the HIV epidemic, turning it from a life-threatening into a manageable chronic disease. This change implies an associated increase in the life expectancy of persons living with HIV, and with this, an increase in age-associated NCDs such as cardiovascular diseases, cancer, and diabetes (15, 16). Moreover, there is a growing concern of the high prevalence of NCDs such as diabetes, obesity, hypertension, and other cardiovascular diseases in both rural and urban African communities (17–19), regardless of HIV serostatus. The WHO predicts that SSA will experience an increase in the prevalence of cardiovascular diseases over the next decade (20), a reasonable prediction considering the rapid epidemiological and nutritional transitions in this region of the world (21). Therefore, multimorbidity is expected to rise and become one of the main drivers of health detriment in African populations. South Africa (SA) is one of the countries with the largest HIV epidemics worldwide, but also has a growing burden of NCDs, potentially facilitating the collision of infectious and non-infectious diseases. However, there is a very limited understanding of the distribution and the co-occurrence of different health conditions in SA. Understanding the magnitude of disease co-occurrence and the nature and type of disease clustering is essential to identify the affected communities and direct the delivery of health services to them. Optimization of health care delivery is crucial, especially in countries with resource-constrained health systems. SA has been largely focused on tackling HIV, TB, and other infectious diseases with relatively few resources allocated to NCDs (22). The implementation of mapping and spatial analyses in the study of chronic diseases has been proven to be an effective methodology for uncovering health determinants and vulnerable populations at higher risk. Despite the potential spatial analysis to advance our understanding of co-occurrent conditions (23), these approaches have focused primarily on single conditions with little research on the complex geography of converging epidemics.

This study aimed to identify spatial patterns emerging from the convergence of different epidemics in vulnerable communities in SA. Using a large nationally representative survey, we conducted a geospatial analysis of the prevalence of HIV, TB, hyperlipidemia, diabetes, cardiovascular and respiratory diseases to identify areas with a high burden of co-occurring diseases. Additionally, we explored the individual risk determinants of co-occurring diseases. Results from this study increase our understanding of disease convergence, support the identification of high-risk subpopulations vulnerable to clustering epidemics, and ultimately may facilitate targeted control interventions that take into account the assembly of co-occurring epidemics.

## METHODS

### Study area and data sources

Data for this study was obtained from the South Africa Demographic and Health Survey (SADHS) conducted in 2016 (24). Briefly, the SADHS is a cross-sectional household survey designed to collect nationally representative data on population, health, and socioeconomic variables (24). Individuals were enrolled in the SADHS via a two-stage sampling procedure to select households. Men and women aged 15-59 in the selected households were eligible for the study. Of 12,132 participants (3,618 men and 8,514 women), 4,862 individuals (2,136 men and 2,726 women) who interviewed and provided specimens for HIV testing were included (24, 25). The global positioning system was used to identify and record the geographical coordinates of each SADHS sample location (26). Figure 1 illustrates the geolocation of each of the sample locations (DHS clusters). Respondents who participated in the testing received educational materials and referrals for free voluntary counseling and testing. HIV serostatus was determined using the enzyme-linked immunosorbent assay (ELISA), based on a parallel testing algorithm. Further details related to the SADHS methodology, study design, and data can be found elsewhere (24, 25).

**Figure. 1.**
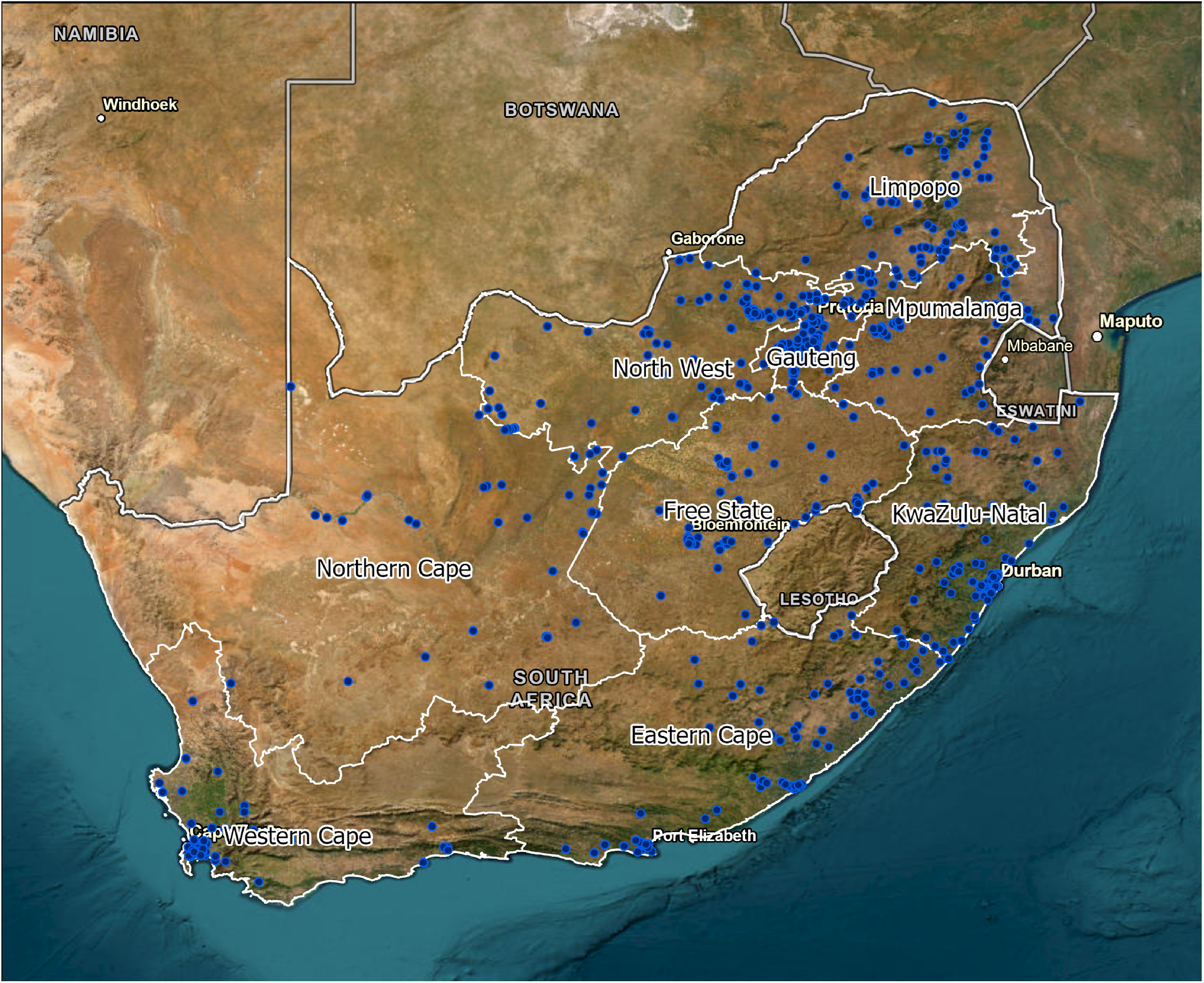
Sample locations. Blue dots illustrating the Demographic and Health Survey sample locations for South Africa Demographic and Health Survey (SADHS) in 2016.

SADHS also collected data for adult morbidity that include the self-reported health status for TB, diabetes, hyperlipidemia, cardiovascular and respiratory diseases. The presence of these diseases was estimated using self-reporting data with the dichotomic ‘yes’ or ‘no’ response to the question, ‘has a doctor, nurse or health worker told you that you have or have had any of the following conditions: TB, diabetes, high blood cholesterol, heart attack or angina/chest pains, stroke, high blood pressure, chronic bronchitis, emphysema, or chronic obstructive pulmonary disease (COPD), asthma?’, Cardiovascular disease was defined as the presence of any of the following conditions: heart attack or angina/chest pains, stroke, or high blood pressure. Respiratory disease was defined as the presence of any of the following conditions: chronic bronchitis, emphysema, COPD, or asthma.

### Geospatial and data visualization analysis

We implemented different spatial and data visualization analyses to evaluate the spatial structure and associations between the six diseases of interest (HIV, TB, diabetes, hyperlipidemia, cardiovascular, and respiratory diseases). First, the estimation of the prevalence for each of the six diseases was spatially smoothed using a kernel smoothing interpolation technique (27). To generate meaningful spatial results for policymaking decisions and comparisons, the spatially smoothed averages were aggregated by district level, which was the spatial resolution used for the remaining of the analysis. The geospatial structure of the prevalence of each disease was identified using optimized hotspot analysis (28, 29). The correlations between diseases were identified estimating Pearson correlation coefficients. The geospatial association between a diseases at the district level was assessed using spatial bivariate and multivariate analysis using the geospatial GeoDa environment (30). For this geospatial analysis, the first step was to identify the spatial correlations between a combination of two diseases using bivariate local indicators of spatial association (LISA). The bivariate LISA statistics identified significant spatial clustering based on the degree of linear association between the prevalence of one disease at a given location and the prevalence of the other disease at neighboring locations (31, 32). Maps were generated illustrating the locations with statistically significant associations and the type of spatial association between the prevalence of both diseases (i.e. high–high, low–low, low–high, and high-low). The second step was to identify the multivariable spatial associations between the distribution of all six diseases combined simultaneously using a K-means clustering analysis, which has been previously used in the study of multivariable clustering of health determinants (33–36). K-means is a partitioning clustering method in which the data are partitioned into k groups (i.e., fourth groups). In this clustering method, the n observations are grouped into k clusters such that the intra-cluster similarity is maximized (or dissimilarity minimized), and the between-cluster similarity minimized (or dissimilarity maximized). A further detailed description of these geospatial methods can be found elsewhere (37, 38). Percentages for the prevalence of each disease in every cluster identified were reported. Maps of the results were generated using ArcGIS Pro (39).

### Individual risk assessment of disease co-occurrence

We assessed the individual odds of co-occurrence of HIV and the other five diseases included in the analysis, controlling for selected sociodemographic and behavioral covariates from the SADHS dataset. The following six socioeconomic and behavioral variables, which have been previously associated with the risk of HIV (40, 41), were included in the analysis: sex, age, poverty estimated using the DHS wealth index, place of residence (urban/rural), education level, and type of union. We fitted survey-weighted multiple logistic regression models with the svyglm() function from the R survey package to account for the two-stage sampling survey design of SADHS, and the adjusted odds ratio (aOR) was calculated by exponentiating the coefficient of multiple logistic regression. We generated two models, in Model 1 the main outcome of the model was HIV serostatus, and for Model 2 we included cardiovascular disease status, the most prevalent chronic health condition identified in this study, as the main outcome.

### Data availability statement

Data are available in a public, open-access repository.

The data that support the findings of this study are available from the Demographic and Health Surveys (http://www.measuredhs.com), but restrictions apply to the availability of these data, which were used under license for the current study and so are not publicly available. However, data are available from the authors on reasonable request and with the permission of Demographic and Health Surveys. We sought and were granted permission to use the core data set for this analysis by Measure DHS.

### Ethics considerations

Procedures and questionnaires for standard Demographic and Health Surveys have been reviewed and approved by the ICF International Institutional Review Board (IRB). The ICF International IRB ensures that the survey complies with the US Department of Health and Human Services regulations for the protection of human subjects, while the host country IRB ensures that the survey complies with the laws and norms of the nation http://dhsprogram.com. Institutional review board approval and informed consent were not necessary for this cross-sectional study because all data were deidentified and publicly available (Common Rule 45 CFR §46). This study follows the Strengthening the Reporting of Observational Studies in Epidemiology (STROBE) reporting guideline (42).

### Funding

Research reported in this publication was supported by the Fogarty International Center (R21 TW011687; D43 TW010543), the National Institute of Mental Health, and the National Institute of Allergy and Infectious Diseases (K24 HL166024; T32 AI007433) of the National Institutes of Health. The contents of this manuscript are solely the responsibility of the authors and do not necessarily represent the official views of the funders.

## RESULTS

All six diseases included in the study exhibited a marked spatial structure with clustering of the diseases located in specific areas within SA (Figure 2). The estimated national spatially smoothed HIV prevalence was 19.%. The eastern region of the country, mostly within the Gauteng, Mpumalanga, and Kwazulu-Natal provinces had a higher HIV prevalence (hotspots; red areas in maps on the left in Figure 2) with a spatially smoothed HIV prevalence of 23.7%. While the southern part of the country, mainly within the Western and Eastern Cape, exhibited the lowest HIV prevalence with a value of 14.7% (coldspots; blue areas in maps on the right in Figure 2). In contrast, the NCDs included in the analysis showed a concentration of the diseases in the center and southern regions of the country. Cardiovascular diseases had the highest smoothed prevalence, with a national average of 22%, followed by respiratory diseases with 4.7% and diabetes with 4.6%. A cardiovascular disease hotspot was identified in the southern part of the country, within the provinces of Free State, and Western and Eastern Cape, with a prevalence of 29.3%, compared to the 16% observed within the coldspots located in the northern and eastern part of the country.

**Figure. 2.**
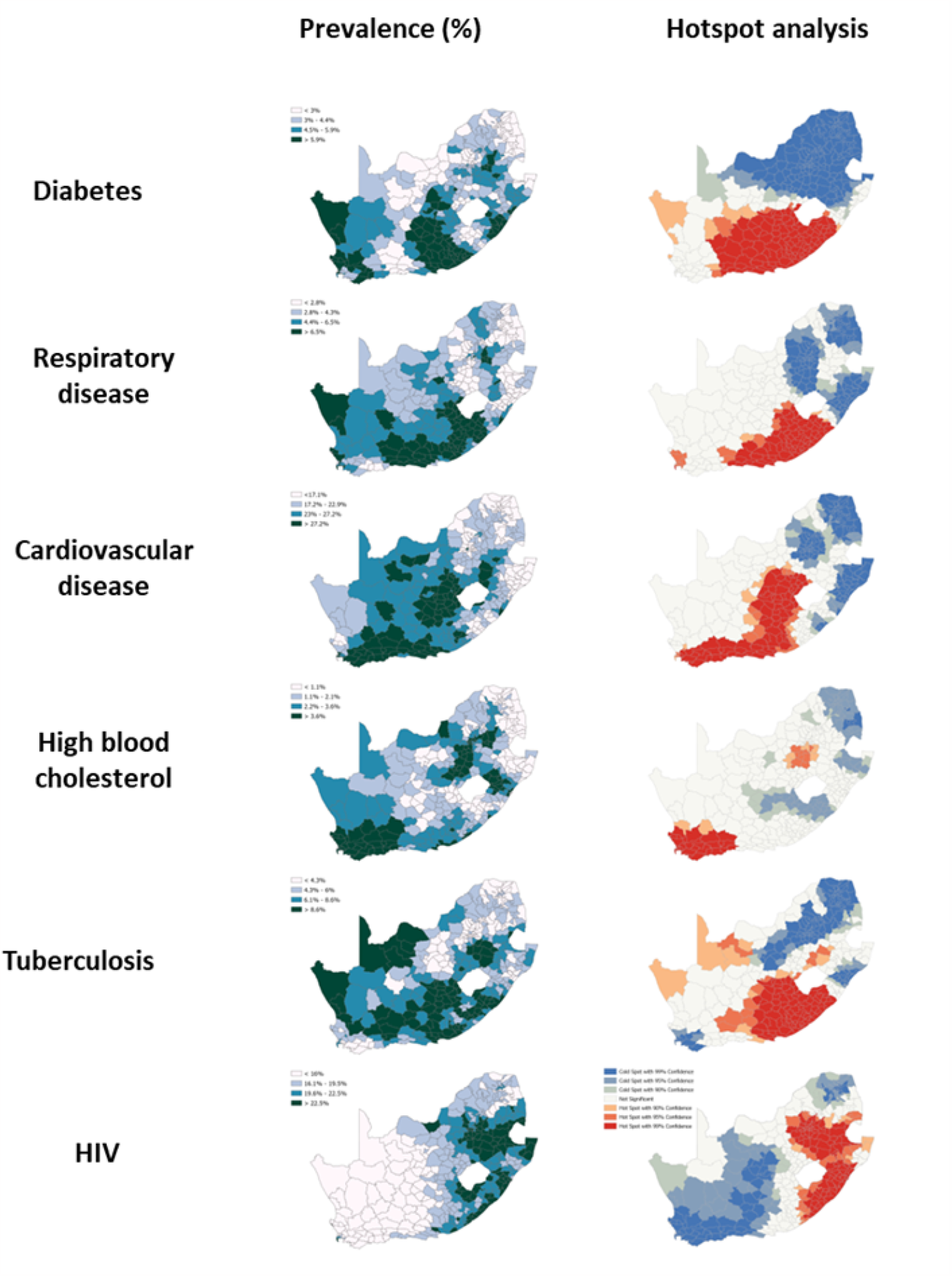
Spatial structure of the prevalence of the six diseases included in the study in South Africa: diabetes, respiratory disease, cardiovascular disease, high blood cholesterol, tuberculosis, and HIV. Maps on the right illustrate the results of the optimized hotspot analysis for each disease, with areas in red illustrating the identified hotspots, whereas areas in blue correspond to the location of the coldspots. White areas illustrate no significant results.

Pearson correlation analyses indicated a lack or negative correlation between district prevalence of HIV and all other five diseases, whereas cardiovascular diseases exhibited positive correlations with all three chronic diseases, diabetes (0.41), respiratory diseases (0.44), and high blood cholesterol (0.44) (Figure 3). These results were consistent with the LISA bivariate analyses and scatterplots that identified a lack of spatial correlation between HIV and the other diseases in most parts of the country (Figure 4). Spatial correlations between cardiovascular diseases with diabetes and respiratory diseases were mainly distributed in the provinces of Free State and Eastern Cape, and with high blood cholesterol within the Western Cape province.

**Figure. 3.**
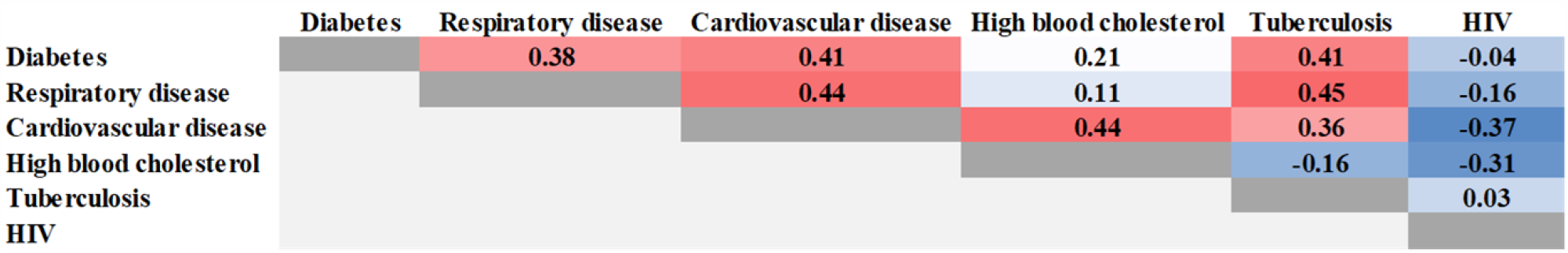
Results from the Pearson correlation analysis of the comparisons between diseases. Red color indicates positive correlation whereas blue color indicates negative correlation between diseases

**Figure. 4.**
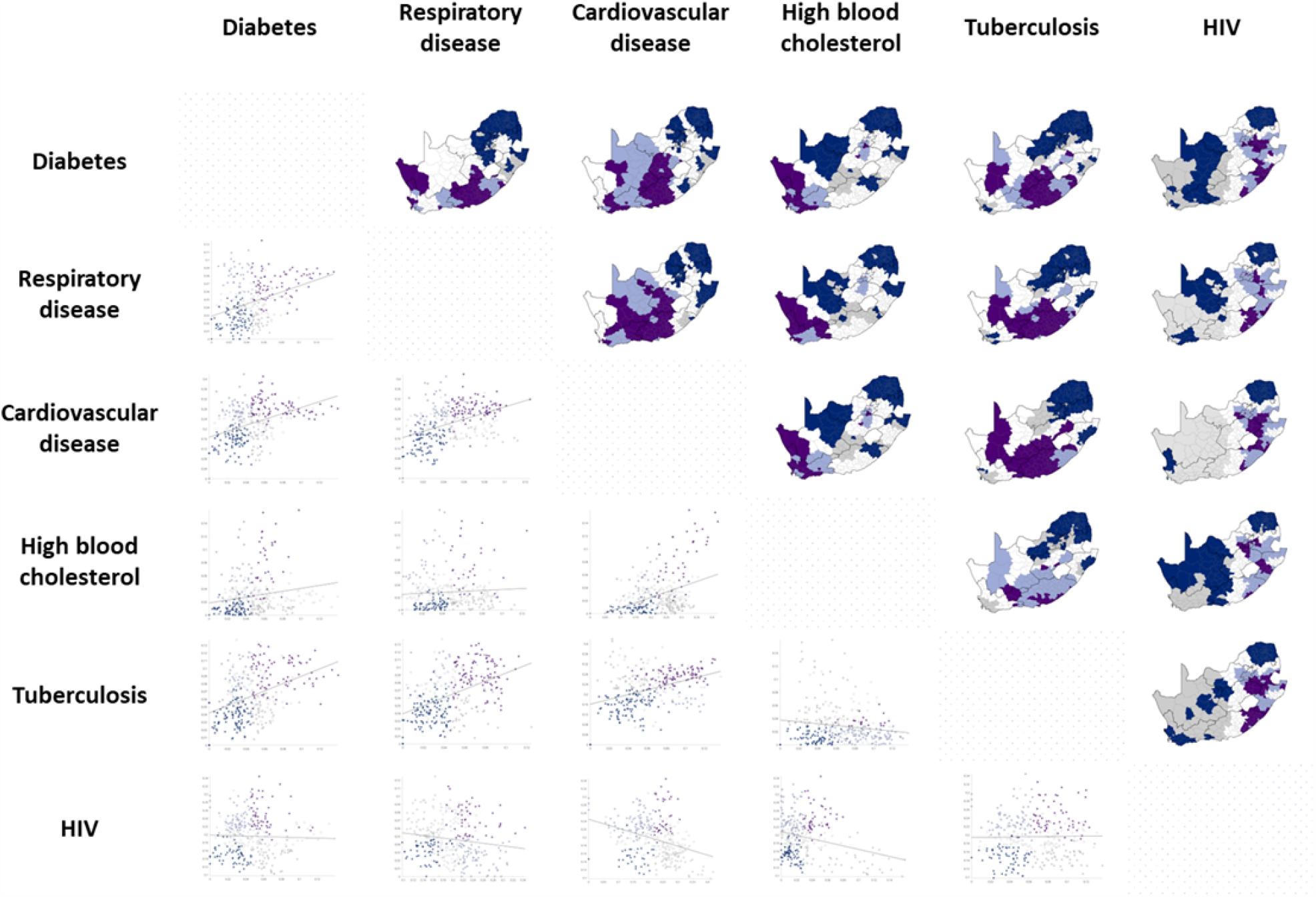
Linear association of spatial autocorrelation (LISA) analysis for the combination between the diseases included in the study. Dark purple color indicates high-high association, dark blue for low-low, light purple for low-high, grey for high-low, and white for nonsignificant association. Scatterplots illustrate the linear association between the prevalence of both diseases at district level.

The multivariable clustering analysis identified five clusters of the combinations among all six diseases (Figure 5). A cluster located within the Western Cape and part of the Northern Cape provinces showed the highest prevalence of cardiovascular diseases (27.4%), high blood cholesterol (7.6%), and the second highest prevalence of diabetes (4.9%) and of respiratory diseases (5.9%) (Cluster 5; areas in dark green color in map in Figure 5). In line with the previous analyses, this Cluster 5 had the lowest HIV prevalence (13.7%). In contrast, Cluster 3 (darker blue areas in map in Figure 5) located mainly in the Kwazulu-Natal province had the highest HIV prevalence (24.3%) and one of the lowest prevalence of cardiovascular diseases (17.8%), high blood cholesterol (2.4%), diabetes (4.4%), and respiratory diseases (2.9%).

**Figure. 5.**
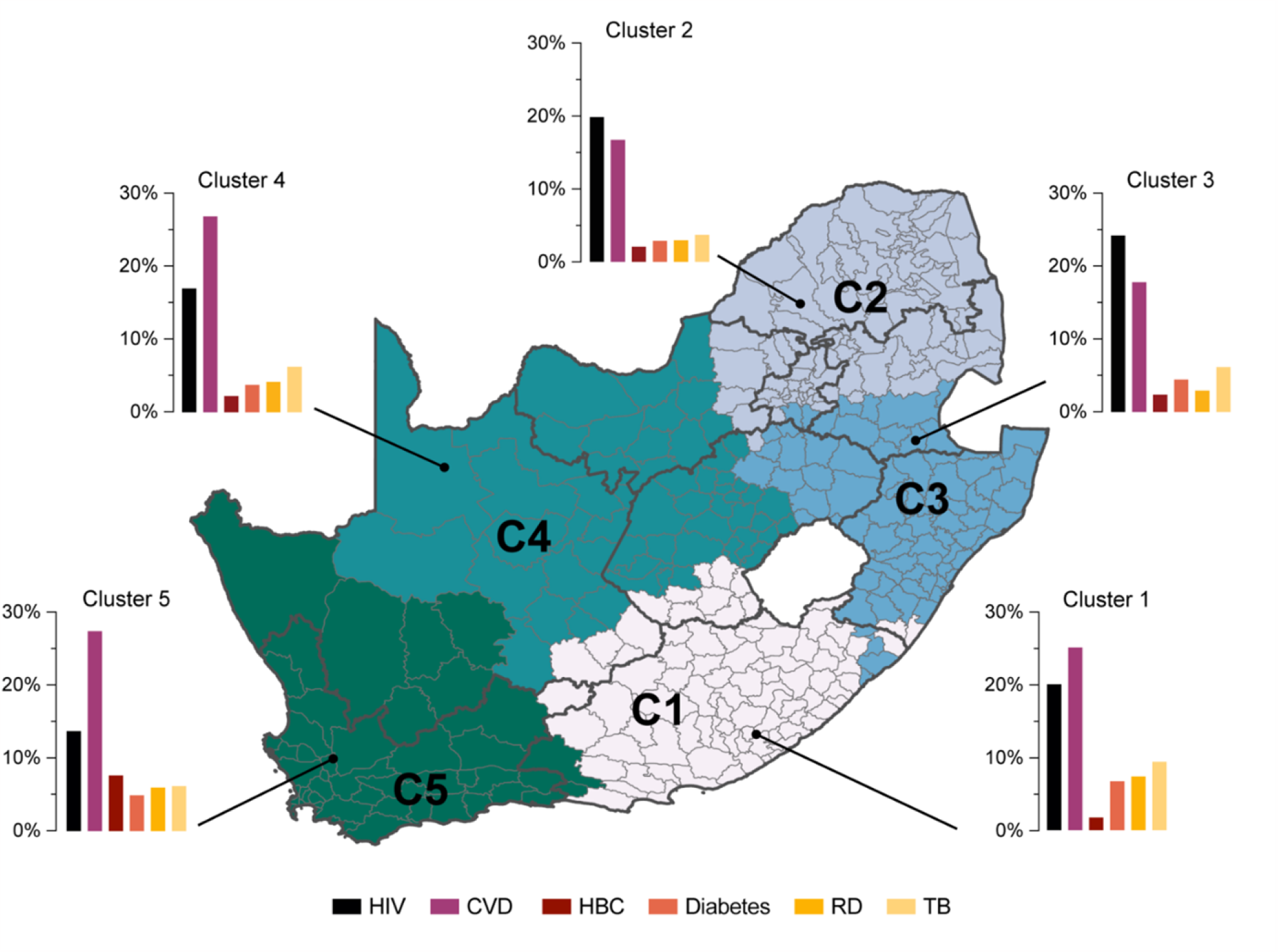
Multivariate K-means clustering analysis for all six diseases combined. Bar plots represent the prevalence for each of the six diseases for the different clusters. CVD: cardiovascular diseases; HBC: high blood cholesterol; RD: respiratory diseases; TB: tuberculosis.

Individual level analyses were consistent with the spatial correlations and found no statistically significant associations between the risk of HIV infection and the presence of noncommunicable diseases included in the study (Figure 6A): cardiovascular diseases (aOR: 0.81; 95% confidence interval [CI]: 0.61-1.09), diabetes (aOR: 0.96; 95% CI: 0.42-2.22), respiratory diseases (aOR: 1.13; 95% CI: 0.65-1.96), and high blood cholesterol (aOR 0.40; 95% CI: 0.13-1.16) after controlling for relevant sociodemographic and behavioral factors. By contrast, the presence of diabetes, respiratory diseases, and high blood cholesterol were significantly associated with an increased risk of cardiovascular diseases (aOR of 5.91, 3.41, and 11.01 respectively; Figure 6B).

**Figure. 6.**
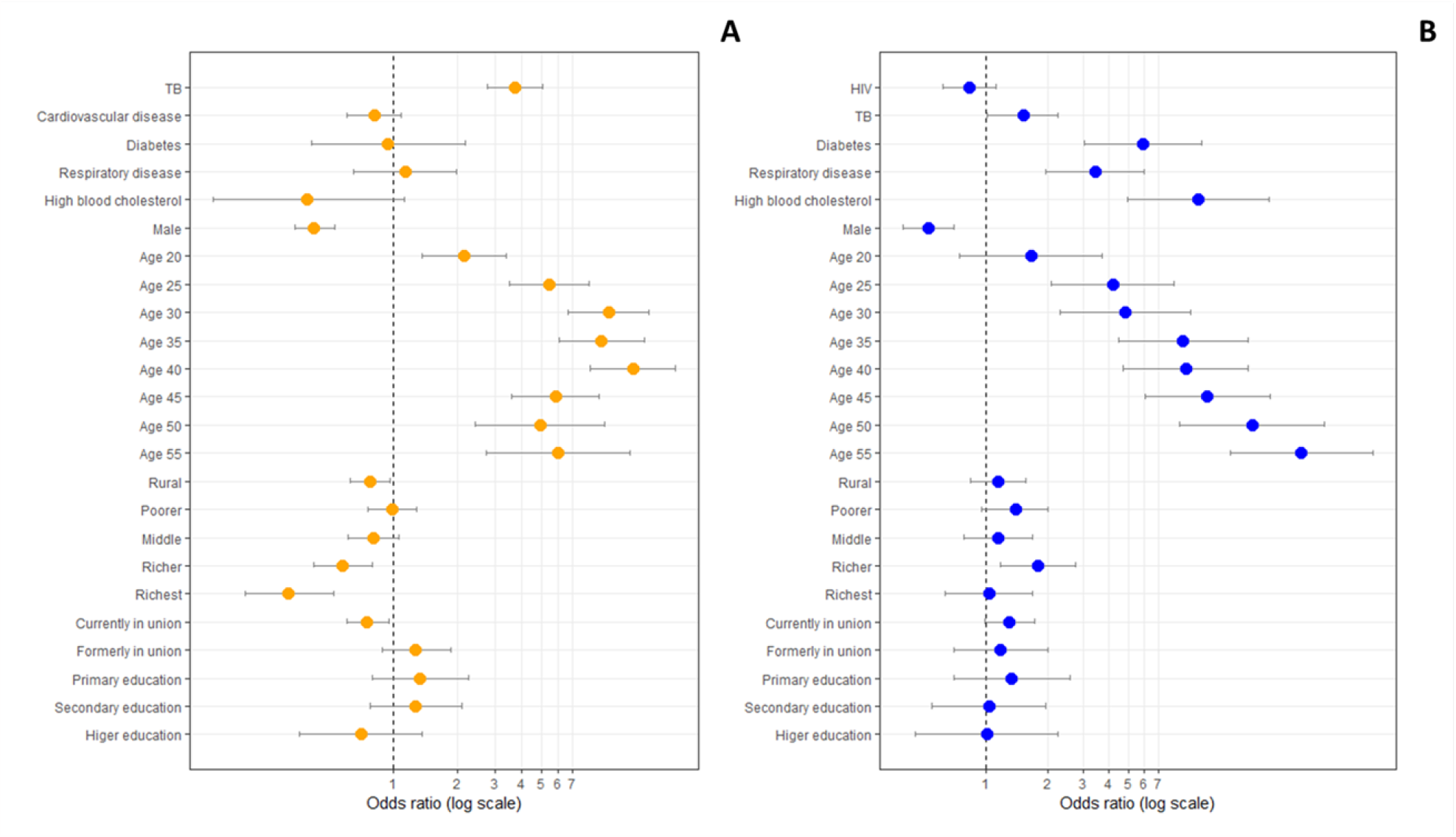
Results of the individual level analysis. In A) Model1, with HIV as the outcome, and B) Model 2, with cardiovascular diseases as the outcome

## DISCUSSION

In this ecological study, we found no evidence of geospatial convergence between the prevalence of HIV and the prevalence of cardiovascular diseases, diabetes, respiratory diseases, and high blood cholesterol in SA. Our results suggest that regions in SA are experiencing different clustered epidemics, with a non-overlapping distribution between HIV and NCDs. Using data from a large national representative survey including individuals age 15-59, we found that the burden of HIV is highest in the eastern part of the country, between the provinces of Gauteng, Kwazulu-Natal, and Mpumalanga. In contrast, the highest burden of NCDs was concentrated in the south central and western part of SA, within the Free State, Eastern and Western Cape. Although they used very different methods, these results are consistent with a previous study conducted in a rural KwaZulu-Natal (43), where the authors found that the geospatial structure of the HIV prevalence had a distinct pattern compared to distribution of other chronic health conditions including elevated blood glucose and elevated blood pressure.

Consistent with the geospatial results, associations at individual level were also characterized by a lack of association between being HIV-seropositive and the presence of any of the NCDs included in the study. Conversely, the presence of cardiovascular diseases was associated with the presence of other NCDs, including diabetes, respiratory diseases, and high blood cholesterol. Similar results were reported by Chang and coworkers in a study conducted in SA (44), in which the authors found a high prevalence of multimorbidity without differences in the likelihood of noncommunicable diseases among persons living with or without HIV. Similarly, a study conducted using the SADHS 2016 found little evidence of association between HIV and increased prevalence of hypertension or diabetes (45). Likewise, a study using data from Demographic and Health surveys conducted in four African countries found no association or negative association between HIV and self-reported hypertension or diabetes in these countries (46). Lastly, a similar study conducted in Zambia and Western Cape found negative association between HIV-seropositive status and diabetes (47).

Collectively, these results illustrate the complex interactions between infectious diseases like HIV and other chronic health conditions. The heterogeneous epidemiological landscapes in SA could be shaped by communities suffering distinct epidemics, possibly driven by the uneven distribution of health and socioeconomic determinants that fueled the infectious and NCDs epidemics at different intensities between regions in the country. Multimorbidity patterns are not characterized merely by co-occurring conditions, but rather exemplify the nature of interactions among social and health determinants that exacerbate the local heterogeneity of the severity or progression of the different health conditions. For example, being the Western cape province, one of the most urbanized in SA, the increase in the incidence of these metabolic risk factors could be associated with accelerated changes in the diet, such as higher consumption of saturated fats, trans fats, cholesterol, and alcohol. In a similar way, the increase in pollution and smoking rates in these urban areas can be significant determinants in the concomitant occurrence of chronic respiratory diseases.

Given that SA continues to be one of the countries with the highest burden of HIV in the world, most of the budget for healthcare is directed towards HIV interventions like antiretroviral therapy. As a result, there is limited funding and capacity to effectively deliver healthcare to those living with other chronic health conditions (48). At the same time social disintegration and inequality, compounded by the weakening economy in the country, have potentially hindered the response to the emerging NCDs epidemics. This undesirable mixture of factors is happening in a health system that is barely coping the high burden of infectious diseases, and that without integrated health management systems and the current surge of NCDs, could experience a serious shortage in healthcare resources. Preventive care and early diagnosis are some of the current interventions to mitigate the incidence of NCDs. Restructuring of the primary healthcare system, especially in those areas with high prevalence of NCDs, is needed for diagnosis and management of patients living with NCDs without affecting the management of the existing HIV and TB epidemics. But difficulties such as underfunding, poor infrastructure, lack of public recognition, and identification of the impact of NCDs will continue to hinder the effective implementation of both community-based health programs targeting communities at higher risk of communicable and non-communicable epidemics emerging in the country. As a result, reforming the infrastructure of the health services delivery to include to provide the needed NCD care based on sustainability and scalability is needed. The first step for this healthcare delivery restructuring is the identification of the uneven distribution of epidemics within the country, and the implementation of geotargeted resource allocation based on the specific epidemiological needs of the affected community. This strategy could also integrate chronic disease management models developed mainly from lessons learnt in the delivery of HIV in the country, in which the robust HIV care platform may improve primary healthcare delivery for other chronic health conditions (49).

It is important to note, however, that the population included in our study is younger than 60 years old. With an aging HIV-positive population, interactions among chronic conditions are likely to emerge. Likewise, distinct epidemic stages might generate a mismatch on the epidemic intensities of the diseases that are colliding in the same communities, in which multimorbidity interactions cannot be captured by general epidemiological measures like disease prevalence. Therefore, studies including more granular measures of the disease stages might be necessary to disentangle disease interactions within communities.

## Limitations

There are several study limitations worth noting. First, some of the variables included in the study could have been affected by inherent biases in the data due to the multiple logistical difficulties in conducting SADHS, such as variability in response rates to HIV testing. The sample weighting procedures were applied to correct for differential non-responses, considering province characteristics, residential place, and sex. We also acknowledge that the validity of self-reported prevalence estimates for NCDs, and particularly those that require access to laboratory and other health infrastructure, can be quite variable (50, 51). Moreover, the displacement process of the SADHS sampling data point via the global positioning system might cause potential spatial bias (41). The methodology used in this analysis based on the geographical location of the SADHS sampling data could have an impact on the precision of the sample location by a few kilometers. Second, chronic health condition statuses for cardiovascular diseases, diabetes, respiratory diseases, and high blood cholesterol were self-reported. Given the high burden of undiagnosed chronic health conditions and variability in screening across regions, this may result in under-reporting in the dataset and thus low estimates in this analysis. Third, our study excludes adults older than 59 years old. This was done to focus on the spatial patterns attributable to the productive age group of 15–59 years, which is the population included in the SADHS 2016. However, it is hereby acknowledged that this limits the ability to evaluate patterns in the age groups that are at the highest risk of chronic health conditions like cardiovascular diseases (60 years and older).

## CONCLUSIONS

We found no evidence of geospatial convergence between the HIV epidemic and other chronic health conditions in a population age 15-59 in SA. By contrast, we found that conditions are distributed in distinct parts of SA, with areas of the highest HIV prevalence clustering in the eastern part of the country, whereas several chronic health conditions like cardiovascular diseases, diabetes, and high blood cholesterol are overlapping in the south and southwestern part of SA. In this study, we identified the geospatial distribution of vulnerable communities with different health needs within SA. A collision of NCDs in the Free State and Western Cape provinces evidenced the need for healthcare expansion in these areas, focusing on improving healthcare services targeting NCDs affecting the communities residing in these provinces. Likewise, the robust HIV service infrastructure implemented during the past few decades to tackle the devastating HIV epidemic in SA can be expanded to provide services for other chronic health conditions, particularly in the areas where communities suffering the highest burden of HIV is concentrated and where NCDs epidemics are likely to emerge in the coming years.

## Data Availability

Data are available in a public, open-access repository. The data that support the findings of this study are available from the Demographic and Health Surveys (http://www.measuredhs.com), but restrictions apply to the availability of these data, which were used under license for the current study and so are not publicly available. However, data are available from the authors on reasonable request and with the permission of Demographic and Health Surveys. We sought and were granted permission to use the core data set for this analysis by Measure DHS.

https://dhsprogram.com/data/

